# Hypertension-Associated Anisometropia: Interocular Choroidal and Macular Asymmetry as Potential Mediators

**DOI:** 10.64898/2026.09.26.26364065

**Authors:** Rongkai Bao, Wurongyue Zhang, Shuilong Lin, Meiying Lin

**Author notes:** Research Project on Chronic Disease Management in 2025 (Grant No: GWJJMB202510025142).

## Abstract

**Purpose:** This study aimed to investigate the association between hypertension and anisometropia and determine whether interocular structural asymmetry mediates this relationship.

**Methods:** In this cross-sectional study, nationally representative samples from the US (N = 12,749; NHANES 1999-2008) and Korea (N = 35,484; KNHANES 2008-2024) were analyzed. Associations between hypertension, blood pressure components (BPC), and anisometropia were examined using weighted multivariable logistic regression and restricted cubic splines. Mediation analysis assessed whether asymmetry in optical coherence tomography (OCT) structural parameters mediated the association between BPC and anisometropia.

**Results:** Hypertensive individuals had higher odds of anisometropia in both cohorts (NHANES: OR = 1.32, 95% CI: 1.12–1.56, P = 0.001; KNHANES: OR = 1.12, 95% CI: 1.05–1.20, P < 0.001). Each 10 mmHg increase in systolic blood pressure (SBP) and pulse pressure (PP) was associated with 5% and 10% greater risk, respectively. Interocular differences in central macular and choroidal thickness partially mediated the association between BPCs and anisometropia. Time-course bulk RNA-seq identified Rock1 as a candidate gene downregulated under hypertensive conditions.

**Conclusions:** Hypertension was associated with anisometropia across two national cohorts. Interocular differences in central macular and choroidal thickness partially mediated this association, suggesting a link between systemic hemodynamic alterations and ocular structural asymmetry.

**Translational Relevance:** These findings suggest that interocular OCT structural asymmetry may serve as a clinically accessible imaging biomarker to identify individuals at increased risk of hypertension-associated anisometropia.

## 1. Introduction

Anisometropia is a type of visual impairment characterized by a difference in the refractive status of the eyes, which is defined as a difference in spherical and cylindrical refractive error between the right and left eyes^1^. It affects approximately 29.62% and 7.10% of the Chinese population^2^ and Hispanic population^3^, respectively, reflecting the effect of genetic background and environmental exposures on refractive development. Chronic disease is associated with visual impairment^4,5^. However, only a few studies have investigated the physiological effects of systemic disease on anisometropia.

Hypertension is a common chronic disease worldwide^6^. Hypertension may be accompanied by retinal vascular alteration^7^ and visual impairment^8,9^. Functional visual impairment relates to the dysregulation of blood vessels^10^. The choroidal thickness of the more myopic eyes was significantly thinner than that of the less myopic eyes in anisometropia patients^11^. However, the connection between hypertension and anisometropia, as well as the mediating effects of ocular microstructure indicators, remains poorly understood. Bridging this knowledge gap may provide new insights into the pathophysiology of anisometropia and highlight how systemic chronic disease may affect ocular conditions.

Using data from the Korea National Health and Nutrition Examination Survey (KNHANES) 2008–2024 and the National Health and Nutrition Examination Survey (NHANES) 1999–2008, the effects of hypertension and BPC on the prevalence of different subtypes of anisometropia were investigated, and whether ocular microstructure changes mediated the relationship between them was examined in this study. Additionally, potential genes that are responsive to hypertension and related changes in ocular structures were assessed to better understand the role of ocular structure-related genes in the pathogenesis of hypertension-related anisometropia.

## 2. Method

### 2.1. Study population

Data for this study were obtained from two cross-sectional surveys: NHANES (1999–2008) and KNHANES (2008–2024)^12,13^. These studies were conducted by the statistical division of the National Centers for Disease Control and Prevention in the United States and Korea, respectively. The study was conducted as per the 1975 Declaration of Helsinki, and participants provided written consent. In the KNHANES VII (2016–2018) and VIII (2019–2021) surveys, detailed optical coherence tomography (OCT) measurements were available for participants examined between 2017 and 2021^14,15^. Information on the study design, sampling methods, and data collection procedures for NHANES and KNHANES is available on their respective official websites (NHANES: https://www.cdc.gov/nchs/nhanes/; KNHANES: https://knhanes.kdca.go.kr/knhanes/).

The exclusion criteria were as follows: (1) lack of data on blood pressure or hypertension status, (2) individuals for whom ophthalmologic measurements were lacking, (3) individuals with a history of ocular diseases or ocular surgery, (4) individuals who were below 18 years old or above 80 years old, (5) individuals with major systemic diseases, and (6) individuals for whom sampling weights were missing. After screening, the final analysis included 12,749 NHANES and 35,484 KNHANES participants (FigureS 1).

### 2.2. Definition of hypertension

A participant was considered to have hypertension if they met any of the following criteria based on the hypertension guidelines published by the American Heart Association in 2017^16^: (1) those who were previously diagnosed with hypertension by a doctor; (2) those who were under medication for hypertension; (3) participants with three consecutive measurements of systolic blood pressure (SBP) ≥ 140 mmHg and/or diastolic blood pressure (DBP) ≥ 90 mmHg on different days.

### 2.3. Definition of anisometropia

Objective refraction data were collected in a non-dilated state using a Nidek ARK-760 autorefractor. The spherical equivalent (SE) difference was calculated using the formula below. Anisometropia was defined as a difference in SE of ≥ 1.0 D between the eyes^17^.

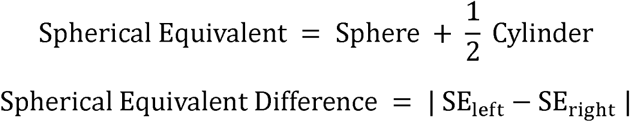

The eye with a greater absolute SE was considered to be the “severe eye”, while the eye with a smaller absolute SE was considered to be the “mild eye”.

Among participants with spherical anisometropia, anisometropia was classified as anisomyopia if either eye was myopic (SE ≤ −1.0 D), or anisohyperopia if either eye was hyperopic (SE ≥ +0.5 D)^18^. Mixed anisometropia was further defined as one myopic eye and one hyperopic eye. Participants with coexisting spherical and astigmatic anisometropia were categorized under spherical anisometropia. Aniso-astigmatism was defined as an interocular difference in refractive astigmatism ≥ 1.0 D^19^.

### 2.4. Covariate assessment

Covariates included demographic characteristics (age, sex, education level, marital status, and poverty-to-income ratio), lifestyle factors (smoking status, alcohol consumption, and physical activity), anthropometric measures (body mass index and body roundness index), and dietary intake variables (energy intake, protein intake, and sodium intake). The use of antihypertensive medication was included as a covariate in analyses where BPC served as the exposure variable. Education level was divided into three categories: less than high school, high school, and more than high school.

### 2.5. Double Machine Learning Framework

To determine the association between hypertension and anisometropia while accounting for high-dimensional and potentially non-linear confounding, the double machine learning (DML) framework^20^ was applied to estimate the average treatment effect (ATE).

We specified a partially linear model of the form:

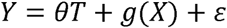

Here, *Y* denotes anisometropia, *T* denotes hypertension status, *X* represents observed confounders, and is the ATE. Two nuisance functions, *E* [*Y* |*X*] and [*T* |*X*], corresponding to the conditional expectations of the outcome and treatment given covariates, were estimated using several machine learning algorithms, including LASSO, random forest, gradient boosting, and support vector regression (SVM). Five-fold cross-fitting was applied to obtain out-of-sample predictions and reduce overfitting bias.

Residuals for hypertension and anisometropia were computed as the differences between observed and predicted values. The ATE was estimated by regressing the outcome residuals on the exposure residuals using ordinary least squares. To evaluate robustness to potential non-linear confounding, the analysis was repeated using models with polynomial-expanded covariates.

### 2.6. Identification of hub gene-regulated hypertension and anisometropia

The bulkRNA-seq data (GSE78042 and GSE302827)^21,22^ were downloaded from the NCBI Gene Expression Omnibus. Hypertension-related genes were obtained from the Comparative Toxicogenomics Database (CTD), DisGeNET, and GeneCards. Differentially expressed genes (DEGs) were identified using the limma-voom pipeline ^23–25^. Genes showing significant condition-time interaction effects were identified and intersected with hypertension-related genes to obtain candidate genes. Time-course expression patterns were further analyzed using ClusterGVis to identify six temporal gene clusters^26^. Next, the Cox regression model was used to identify prognostic genes within Cluster 5 that exhibited the highest number of hypertension-related genes. Group differences in gene expression between HTN and control were assessed by conducting Student’s t-tests (Arhgap6, Fat1, Kalrn, Pam, and Rock1) or the Wilcoxon test (Jak2, Npr3).

### 2.7. Statistical Analysis

Between-group comparisons for continuous variables were conducted using weighted linear regression models, while the weighted Chi-Square test was conducted for categorical variables. For paired ocular comparisons within each anisometropia subtype, the differences between normally distributed data were determined by conducting paired t-tests, whereas the differences between non-normally distributed data were determined by conducting the Wilcoxon signed-rank test; data normality was assessed by the Shapiro-Wilk test.

To investigate the association between the prevalence of anisometropia and hypertension, weighted multivariate logistic regression was performed. Model 1 was unadjusted; Model 2 was adjusted for age and sex; Model 3 was further adjusted for all available covariates.

To evaluate the potential dose-response relationship (linear or non-linear) between BPC and the prevalence of anisometropia, weighted multivariate restricted cubic spline (RCS) regression was applied. This method was used to assess the nonlinearity of the association between BPC and anisometropia.

The ‘mediation’ package in the R software was used for mediation analysis based on bootstrapping calculations to investigate the indirect effect mediated by ocular microstructure and the total effect of BPC on the risk of anisometropia. The proportion of the effect mediated by ocular microstructure was calculated using the formula (mediated effect/total effect) × 100%. Two hypothesized pathways were evaluated: (1) mediation via absolute inter-ocular differences in ocular structure and (2) mediation via relative inter-ocular differences in ocular structure.

For the first hypothesized pathway, the ocular differences in central macular thickness (CMT), subfoveal choroidal thickness (SFCT), axial length (AL), and retinal thickness (RT) were calculated. Next, mediation analysis was conducted to examine whether interocular differences mediated the association between BPCs and the risk of anisometropia. For the second hypothesized mediated pathway, the associations between BPCs and ocular microstructural parameters were first hypothesized using generalized estimating equation (GEE) models. Only BPC-ocular microstructure (BPC-OCT) pairs showing significant associations were retained for subsequent mediation analyses. Next, whether these ocular microstructures mediated the associations between BPCs and the risk of anisometropia was evaluated. Additionally, whether the mediation effects differed between severe and mild eyes was investigated. Moderated mediation analyses were conducted using the ‘manymome’ package in the R software^27^. Each participant contributed two observations in a long-format dataset, with eye severity (severe = 0, mild = 1) serving as the binary moderator. Conditional indirect effects at each level of eye severity were estimated with 2,000 bootstrap resamples (95% CI). The Index of Moderated Mediation^28^, quantifying the difference in indirect between severe and mild eyes, was considered to be statistically significant when its CI excluded zero.

To address missing data, Generative Adversarial Imputation Nets (GAIN)^29^ were applied; this is a deep learning-based imputation method that uses an adversarial training framework to generate imputations consistent with the observed data distribution, as described in another study^30^. All statistical analyses were conducted using R v.4.4.2 and Python v.3.13.5. All results were considered to be statistically significant at P < 0.05 (two-sided).

## 3. Results

### 3.1. Baseline characteristics of participants

This study included 48,233 participants, among which 12,749 participants were from NHANES, and 35,484 participants were from KNHANES, representing 123.33 million and 16.46 million non-institutionalized civilians from America and Korea, respectively (FigureS 1). In the NHANES cohort, 3,599 participants (28.23%) had hypertension, with a mean age of 51.70 years (p < 0.001), and 48.90% of participants were men (p = 0.36). In the KNHANES cohort, 12,908 of 35,484 participants had hypertension. The mean age of the participants with hypertension was 48.63 years (p < 0.001), and 54.80% of participants were men (p < 0.001).

The clinical characteristics of the participants according to hypertension as a column-stratified variable are shown in Table 1. Participants with hypertension tended to be older, male, married, and with college-level education.

In the OCT subcohort, SFCT, RT, and AL were compared between the severe and mild eyes across anisometropia subtypes. For SFCT, significant inter-eye differences were found in anisometropia (275.28 vs. 288.72 μm, P < 0.001, N = 994) and anisomyopia (238.99 vs. 278.94 μm, P < 0.001, N = 342), whereas no significant inter-eye differences were found in anisohyperopia (294.98 vs. 291.43 μm, P = 0.511, N = 167) or aniso-astigmatism (294.09 vs. 294.69 μm, P = 0.811, N = 485). Similarly, RT was significantly lower in the severe eye among anisometropia (276.54 vs. 277.43 μm, P = 0.013, N = 941) and anisomyopia patients (274.87 vs. 277.16 μm, P < 0.001, N = 337), but not in anisohyperopia (276.77 vs. 277.14 μm, P = 0.888, N = 163) or aniso-astigmatism patients (277.74 vs. 277.75 μm, P = 0.811, N = 441). For AL, significant inter-eye differences were observed in anisometropia (24.57 vs. 24.27 mm, P < 0.001, N = 891), anisomyopia (25.84 vs. 25.05 mm, P < 0.001, N = 319), aniso-astigmatism (24.10 vs. 24.04 mm, P < 0.001, N = 420), and anisohyperopia (23.18 vs. 23.26 mm, P = 0.002, N = 156).

**Table 1.**
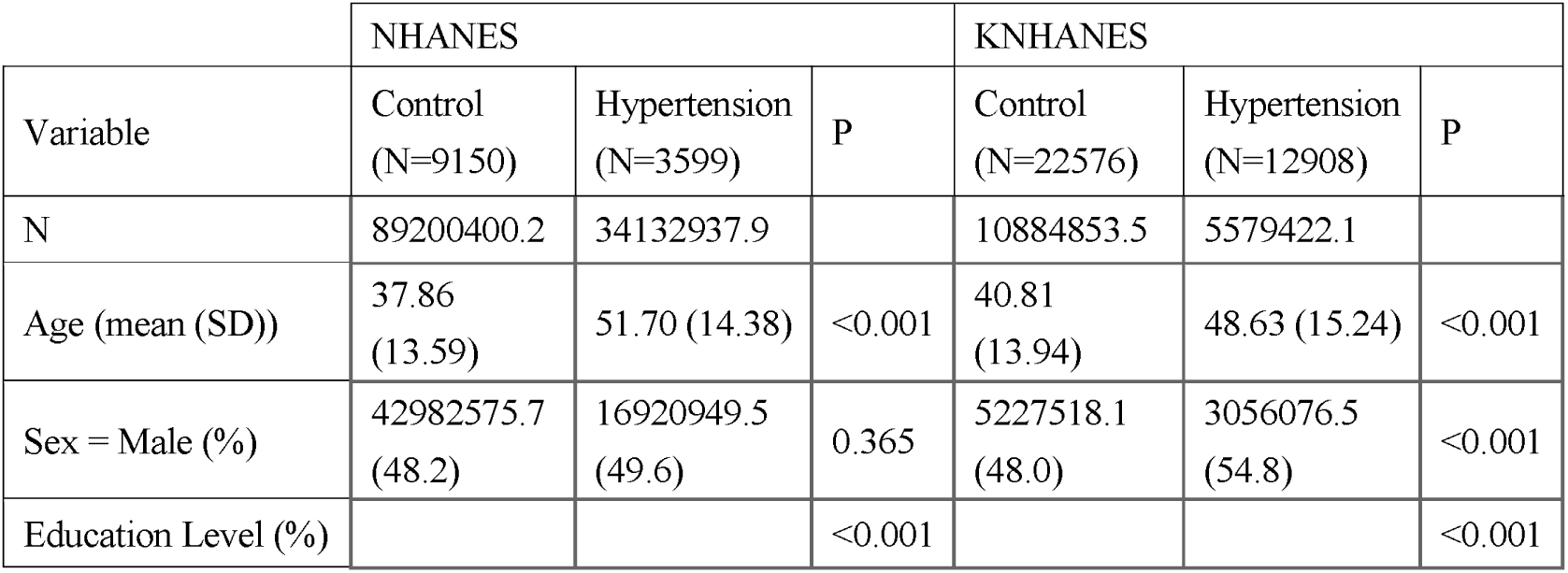
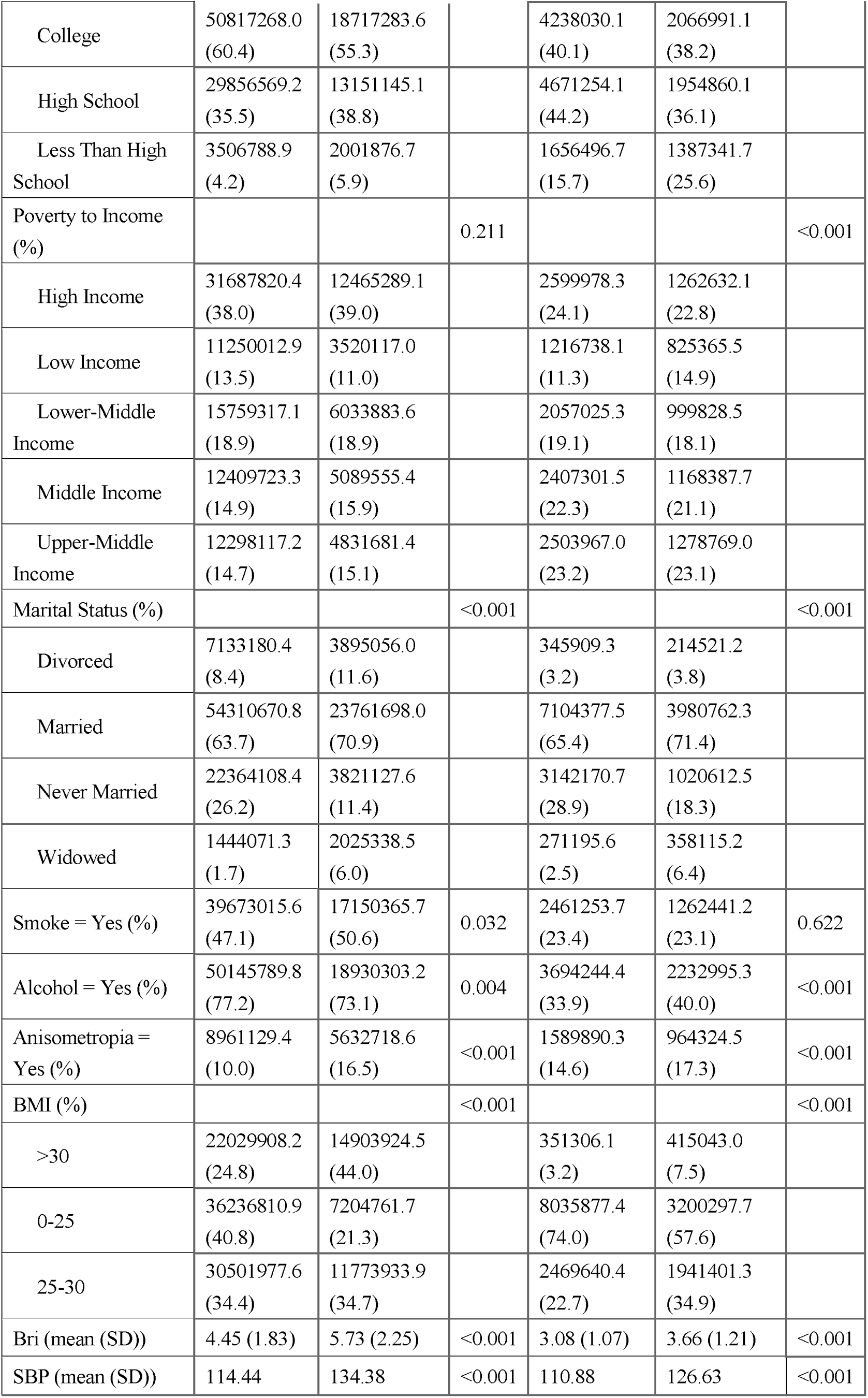
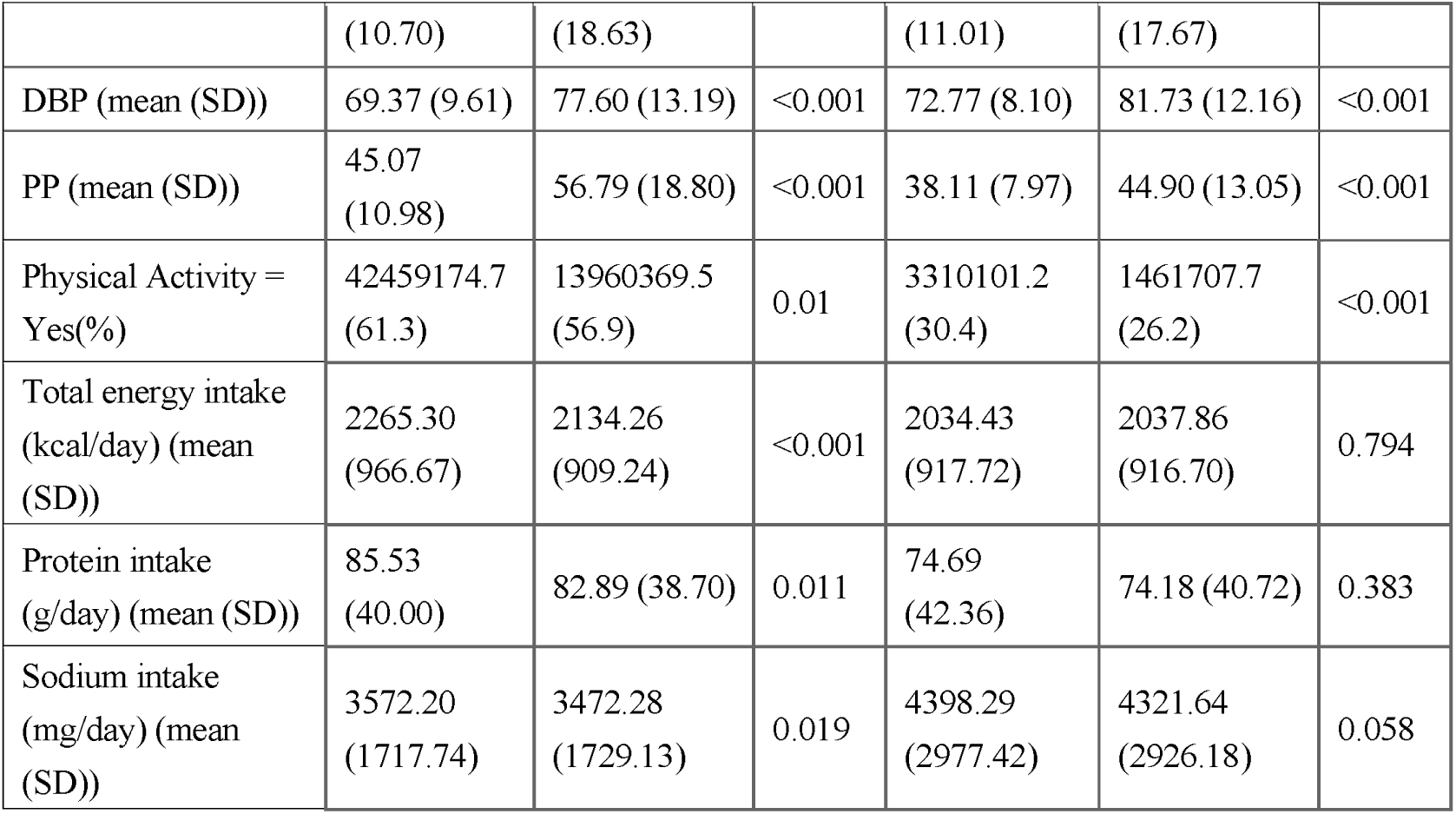
Demographic and clinical characteristics of the participants with and without hypertension. OCT subcohort, N: 4,913–6,839.

| Variable | NHANES |  |  | KNHANES |  |  |
| --- | --- | --- | --- | --- | --- | --- |
|  | Control<br>(N=9150) | Hypertension<br>(N=3599) | P | Control<br>(N=22576) | Hypertension<br>(N=12908) | P |
| N | 89200400.2 | 34132937.9 |  | 10884853.5 | 5579422.1 |  |
| Age (mean (SD)) | 37.86<br>(13.59) | 51.70 (14.38) | <0.001 | 40.81<br>(13.94) | 48.63 (15.24) | <0.001 |
| Sex = Male (%) | 42982575.7<br>(48.2) | 16920949.5<br>(49.6) | 0.365 | 5227518.1<br>(48.0) | 3056076.5<br>(54.8) | <0.001 |
| Education Level (%) |  |  | <0.001 |  |  | <0.001 |
| College | 50817268.0<br>(60.4) | 18717283.6<br>(55.3) |  | 4238030.1<br>(40.1) | 2066991.1<br>(38.2) |  |
| High School | 29856569.2<br>(35.5) | 13151145.1<br>(38.8) |  | 4671254.1<br>(44.2) | 1954860.1<br>(36.1) |  |
| Less Than High School | 3506788.9<br>(4.2) | 2001876.7<br>(5.9) |  | 1656496.7<br>(15.7) | 1387341.7<br>(25.6) |  |
| Poverty to Income (%) |  |  | 0.211 |  |  | <0.001 |
| High Income | 31687820.4<br>(38.0) | 12465289.1<br>(39.0) |  | 2599978.3<br>(24.1) | 1262632.1<br>(22.8) |  |
| Low Income | 11250012.9<br>(13.5) | 3520117.0<br>(11.0) |  | 1216738.1<br>(11.3) | 825365.5<br>(14.9) |  |
| Lower-Middle Income | 15759317.1<br>(18.9) | 6033883.6<br>(18.9) |  | 2057025.3<br>(19.1) | 999828.5<br>(18.1) |  |
| Middle Income | 12409723.3<br>(14.9) | 5089555.4<br>(15.9) |  | 2407301.5<br>(22.3) | 1168387.7<br>(21.1) |  |
| Upper-Middle Income | 12298117.2<br>(14.7) | 4831681.4<br>(15.1) |  | 2503967.0<br>(23.2) | 1278769.0<br>(23.1) |  |
| Marital Status (%) |  |  | <0.001 |  |  | <0.001 |
| Divorced | 7133180.4<br>(8.4) | 3895056.0<br>(11.6) |  | 345909.3<br>(3.2) | 214521.2<br>(3.8) |  |
| Married | 54310670.8<br>(63.7) | 23761698.0<br>(70.9) |  | 7104377.5<br>(65.4) | 3980762.3<br>(71.4) |  |
| Never Married | 22364108.4<br>(26.2) | 3821127.6<br>(11.4) |  | 3142170.7<br>(28.9) | 1020612.5<br>(18.3) |  |
| Widowed | 1444071.3<br>(1.7) | 2025338.5<br>(6.0) |  | 271195.6<br>(2.5) | 358115.2<br>(6.4) |  |
| Smoke = Yes (%) | 39673015.6<br>(47.1) | 17150365.7<br>(50.6) | 0.032 | 2461253.7<br>(23.4) | 1262441.2<br>(23.1) | 0.622 |
| Alcohol = Yes (%) | 50145789.8<br>(77.2) | 18930303.2<br>(73.1) | 0.004 | 3694244.4<br>(33.9) | 2232995.3<br>(40.0) | <0.001 |
| Anisometropia = Yes (%) | 8961129.4<br>(10.0) | 5632718.6<br>(16.5) | <0.001 | 1589890.3<br>(14.6) | 964324.5<br>(17.3) | <0.001 |
| BMI (%) |  |  | <0.001 |  |  | <0.001 |
| >30 | 22029908.2<br>(24.8) | 14903924.5<br>(44.0) |  | 351306.1<br>(3.2) | 415043.0<br>(7.5) |  |
| 0-25 | 36236810.9<br>(40.8) | 7204761.7<br>(21.3) |  | 8035877.4<br>(74.0) | 3200297.7<br>(57.6) |  |
| 25-30 | 30501977.6<br>(34.4) | 11773933.9<br>(34.7) |  | 2469640.4<br>(22.7) | 1941401.3<br>(34.9) |  |
| Bri (mean (SD)) | 4.45 (1.83) | 5.73 (2.25) | <0.001 | 3.08 (1.07) | 3.66 (1.21) | <0.001 |
| SBP (mean (SD)) | 114.44 | 134.38 | <0.001 | 110.88 | 126.63 | <0.001 |
|  | (10.70) | (18.63) |  | (11.01) | (17.67) |  |
| DBP (mean (SD)) | 69.37 (9.61) | 77.60 (13.19) | <0.001 | 72.77 (8.10) | 81.73 (12.16) | <0.001 |
| PP (mean (SD)) | 45.07<br>(10.98) | 56.79 (18.80) | <0.001 | 38.11 (7.97) | 44.90 (13.05) | <0.001 |
| Physical Activity =<br>Yes(%) | 42459174.7<br>(61.3) | 13960369.5<br>(56.9) | 0.01 | 3310101.2<br>(30.4) | 1461707.7<br>(26.2) | <0.001 |
| Total energy intake<br>(kcal/day) (mean<br>(SD)) | 2265.30<br>(966.67) | 2134.26<br>(909.24) | <0.001 | 2034.43<br>(917.72) | 2037.86<br>(916.70) | 0.794 |
| Protein intake<br>(g/day) (mean (SD)) | 85.53<br>(40.00) | 82.89 (38.70) | 0.011 | 74.69<br>(42.36) | 74.18 (40.72) | 0.383 |
| Sodium intake<br>(mg/day) (mean<br>(SD)) | 3572.20<br>(1717.74) | 3472.28<br>(1729.13) | 0.019 | 4398.29<br>(2977.42) | 4321.64<br>(2926.18) | 0.058 |

Before the data were analyzed, missing data were handled using GAIN-based imputation. The imputation model converged rapidly across iterations in both cohorts. The final RMSE across the three iterations was 0.074 ± 0.001 for NHANES and 0.066 ± 0.002 for KNHANES, indicating acceptable imputation performance before subsequent analyses were performed.

### 3.2. The association between anisometropia and the prevalence of hypertension

As shown in Figure 1B, the overall age-standard prevalence of anisometropia in the study was 11.34% (SE = 0.43) and 15.83% (SE = 0.24) in the NHANES and KNHANES cohorts. In both cohorts, individuals with hypertension had a significantly higher prevalence of anisometropia than those without hypertension (NHANES: 14.22% vs. 10.30%; KNHANES: 17.16% vs. 15.06%; both P < 0.001).

**Figure 1:**
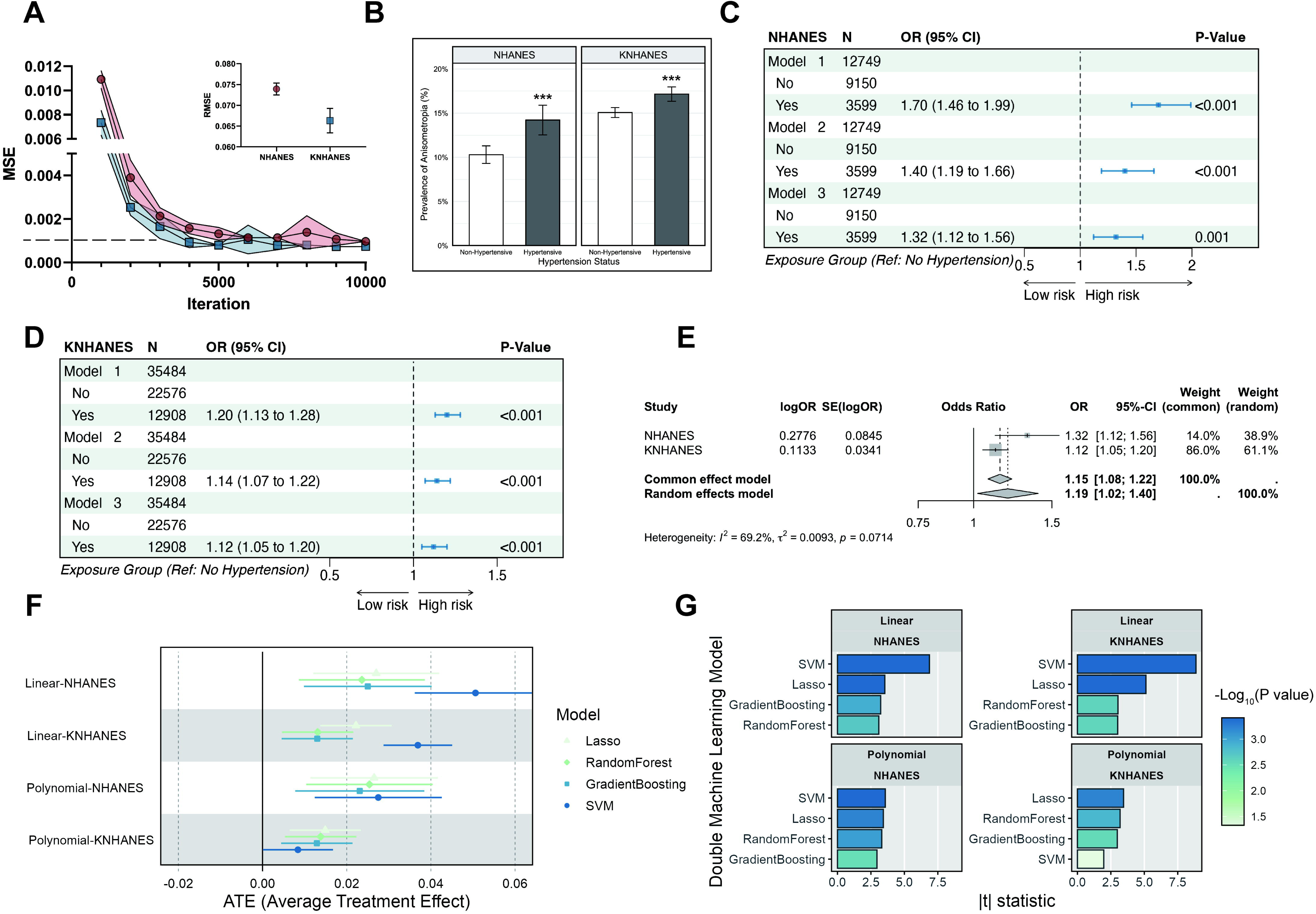
Association between risks of anisometropia and hypertension. (A) The incomplete data were filled using GAIN. (B) The prevalence of anisometropia in two distinct cohorts is illustrated; ***P < 0.001. (C and D) The results of weighted multivariate logistic regression of hypertension and anisometropia in the NHANES and KNHANES cohorts are presented. Model 1: crude model. Model 2: adjusted for age and sex. Model 3: adjusted for all covariates. (E) Meta-analysis of the two cohorts was performed. (F) Forest plot of ATE with 95% CI calculated by the Linear- and Polynomial feature expansion-DML model across different machine learning algorithms (LASSO, Random Forest, Gradient Boosting, and Support Vector Machine) in the NHANES and KNHANES cohorts. (G) The bar plots illustrate the statistical significance of the estimated causal effects, expressed as -log10(P value), for each DML model. The color intensity of the bars represents the absolute t-statistic, with darker colors indicating stronger statistical evidence.

The results of weighted multivariate logistic regression analysis demonstrated robust associations between hypertension and anisometropia across all three models. In the NHANES cohort, hypertension was associated with a significantly increased risk of anisometropia in the unadjusted model (OR = 1.70; 95% CI: 1.46–1.99; P < 0.01). After the covariates were adjusted, a significant correlation between the prevalence of anisometropia and hypertension was observed (Model 2: OR = 1.40; 95% CI: 1.19–1.66; P < 0.001; Model 3: OR = 1.32; 95% CI: 1.12–1.56; P = 0.001), with the association remaining significant after all covariates were adjusted. In the KNHANES cohort, hypertension was similarly associated with a greater risk of anisometropia across all models (Model 1: OR = 1.20; 95% CI: 1.13–1.28; P < 0.001; Model 2: OR = 1.14; 95% CI: 1.07–1.22; P < 0.001; Model 3: OR = 1.12; 95% CI: 1.05–1.20; P < 0.001). To synthesize effect estimates and improve statistical power, meta-analyses were conducted across the two cohorts. The results revealed substantial heterogeneity (I^2^ = 69.2%, p = 0.07), and a random-effects model yielded a pooled OR of 1.19 (95% CI: 1.02–1.40), indicating significantly greater odds of anisometropia among hypertensive individuals.

To determine the effects of hypertension on anisometropia, the ATE was estimated using DML, adjusting for a comprehensive set of confounding factors. The DML analysis confirmed a positive association between hypertension and anisometropia across both cohorts and all machine learning specifications (Figures 1F and G). In the NHANES cohort, ATE estimates ranged from 0.013 to 0.051 (all P < 0.01), and in the KNHANES cohort, the estimates ranged from 0.008 to 0.036 (all P < 0.05), consistent with a robust positive effect across both populations.

### 3.3. The relationship between blood pressure components and the prevalence of anisometropia

The associations between BPCs and the risk of anisometropia were examined using weighted multivariate logistic regression and a random-effects meta-analytic model (Table 2).

**Table 2.** Association between BPCs (SBP, DBP, and PP) and risk of anisometropia.

| Exposure Model |  | NHANES OR<br>(95% CI) | KNHANES OR<br>(95% CI) | Pooled OR (95% CI)* | P<br>(Overall) | P<br>(Heterogeneity) |
| --- | --- | --- | --- | --- | --- | --- |
| SBP | Model 1 | 1.14 (1.09-1.18) | 1.08 (1.06-1.1) | 1.11 (1.05-1.17) | <0.001 | 0.015 |
|  | Model 2 | 1.07 (1.02-1.12) | 1.06 (1.04-1.08) | 1.06 (1.04-1.08) | <0.001 | 0.715 |
|  | Model 3 | 1.05 (1.01-1.1) | 1.05 (1.03-1.07) | 1.05 (1.03-1.07) | <0.001 | 1.000 |
| DBP | Model 1 | 1.06 (0.99-1.14) | 0.98 (0.95-1.01) | 1.01 (0.94-1.09) | 0.750 | 0.045 |
|  | Model 2 | 1.02 (0.95-1.09) | 0.95 (0.92-0.98) | 0.98 (0.91-1.05) | 0.518 | 0.065 |
|  | Model 3 | 1.01 (0.94-1.08) | 0.96 (0.93-0.99) | 0.98 (0.93-1.02) | 0.280 | 0.191 |
| PP | Model 1 | 1.13 (1.09-1.17) | 1.19 (1.16-1.22) | 1.16 (1.10-1.22) | <0.001 | 0.020 |
|  | Model 2 | 1.07 (1.03-1.11) | 1.17 (1.14-1.21) | 1.12 (1.03-1.22) | 0.011 | <0.001 |
|  | Model 3 | 1.06 (1.01-1.1) | 1.15 (1.11-1.18) | 1.11 (1.02-1.20) | 0.014 | 0.002 |

\* Meta-analysis was conducted using the random-effects model (REML).

For SBP, a 10 mmHg increase in SBP is associated with a 5% increase in the risk of anisometropia (pooled OR: 1.05; 95% CI: 1.03–1.07; P < 0.001) as indicated by Model 3. A substantial degree of heterogeneity was observed for pulse pressure (PP) (I^2^ = 91.7%). Using the random-effects model, PP was found to be associated with the risk of anisometropia. The risk of anisometropia increased by 10% for every 10 mmHg rise in PP, as indicated by Model 3 (pooled OR: 1.10; 95% CI: 1.01–1.19; P = 0.027). In contrast, DBP exhibited no significant correlation in the pooled analysis for Model 3 (Pooled OR: 0.98; 95% CI: 0.92–1.02; P = 0.191).

### 3.4. Non-linear relationship between blood pressure components and the anisometropia risk

Weighted RCS analysis was performed to investigate the dose-response relationship between BPCs and the risk of anisometropia (Figure 2). After adjusting for all covariates as in Model 3 plus blood pressure medication, the results revealed significant non-linear associations between BPCs and the risk of anisometropia in the KNHANES and NHANES cohorts (all P-nonlinear < 0.001, except for DBP in the KNHANES cohort).

**Figure 2:**
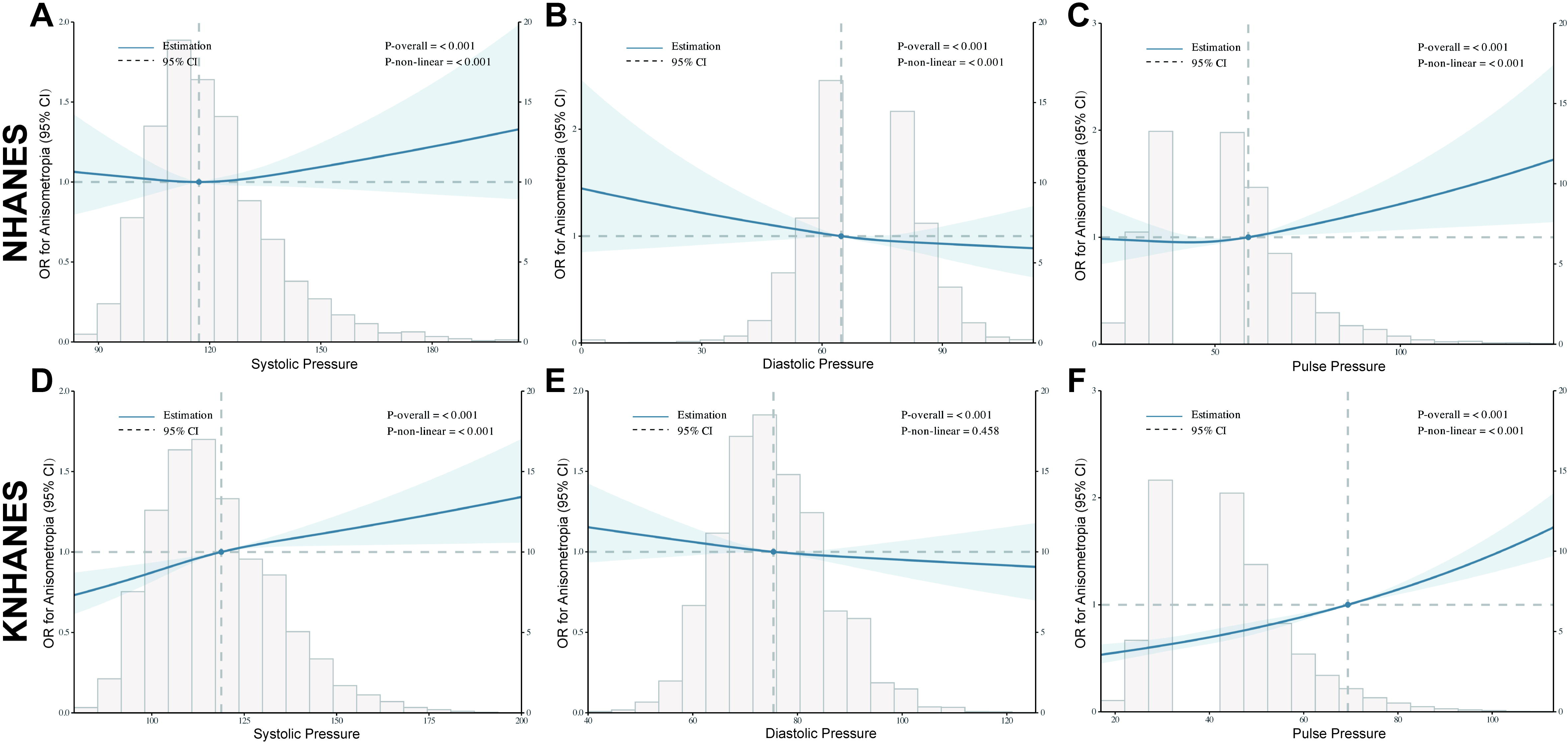
Weighted restricted cubic spline (RCS) curves for the association between BPCs and the risk of anisometropia among the NHANES cohort and the KNHANES cohort. The models were adjusted for all covariates in Model 3 plus blood pressure medication. (A-C) The NHANES cohort; (D-F) The KNHANES cohort. The solid blue lines indicate estimated associations, and the shaded areas represent 95% CIs. RCS: restricted cubic spline; OR: odds ratio.

The risk profiles for SBP and PP generally followed a J-shaped or upward non-linear trajectory, while DBP showed a predominantly L-shaped pattern. For SBP, the two cohorts showed distinct non-linear dynamics. In the NHANES cohort, high SBP, defined as greater than 117.10 mmHg, was associated with a substantial increase in the prevalence of anisometropia. In the KNHANES cohort, high SBP, defined as greater than 118.79 mmHg, was associated with a slight increase in the prevalence of anisometropia.

For PP, a threshold effect was observed in both cohorts: the risk of anisometropia increased steadily below 58.99 mmHg (NHANES) and 69.38 mmHg (KNHANES), and then increased rapidly above these thresholds, forming a J-shaped non-linear pattern. For DBP, the risk of anisometropia decreased sharply at lower diastolic levels, then plateaued beyond 64.78 mmHg in the NHANES cohort and 75.42 mmHg in the KNHANES cohort.

### 3.5. Subgroup analysis and subtype analysis

The results of the subtype analyses, stratified by anisometropia subtypes, revealed differential associations with hypertension and BPC (Figure 3A). The overall age-standard prevalence of aniso-astigmatism, anisomyopia, and anisohyperopia were 7.49%, 2.84%, and 1.38% in the NHANES cohort, and 8.97%, 5.16%, and 0.87% in the KNHANES cohort, respectively. For aniso-astigmatism, SBP was associated with a 5% increase in risk (pooled OR: 1.05 per 10 mmHg increase; 95% CI: 1.03–1.08; P < 0.001), with additional significant associations observed for PP (pooled OR: 1.10 per 10 mmHg increase; 95% CI: 1.01–1.19; P = 0.024). For anisomyopia, hypertension was associated with a 19% increase in risk (pooled OR: 1.19; 95% CI: 1.04-1.36; P = 0.009), and each 10 mmHg increase in SBP was associated with a 4% increase in risk (pooled OR: 1.04; 95% CI: 1.00–1.07; P = 0.049), with PP showing a borderline significant association in the same direction (pooled OR: 1.05; 95% CI: 1.00–1.11; P = 0.051). For anisohyperopia, each 10 mmHg increase in PP was associated with a 12% increase in risk (pooled OR: 1.12; 95% CI: 1.05–1.20; P < 0.001), while each 10 mmHg increase in DBP was associated with a 12% decrease in risk (pooled OR: 0.89; 95% CI: 0.79–0.99; P = 0.028).

**Figure 3:**
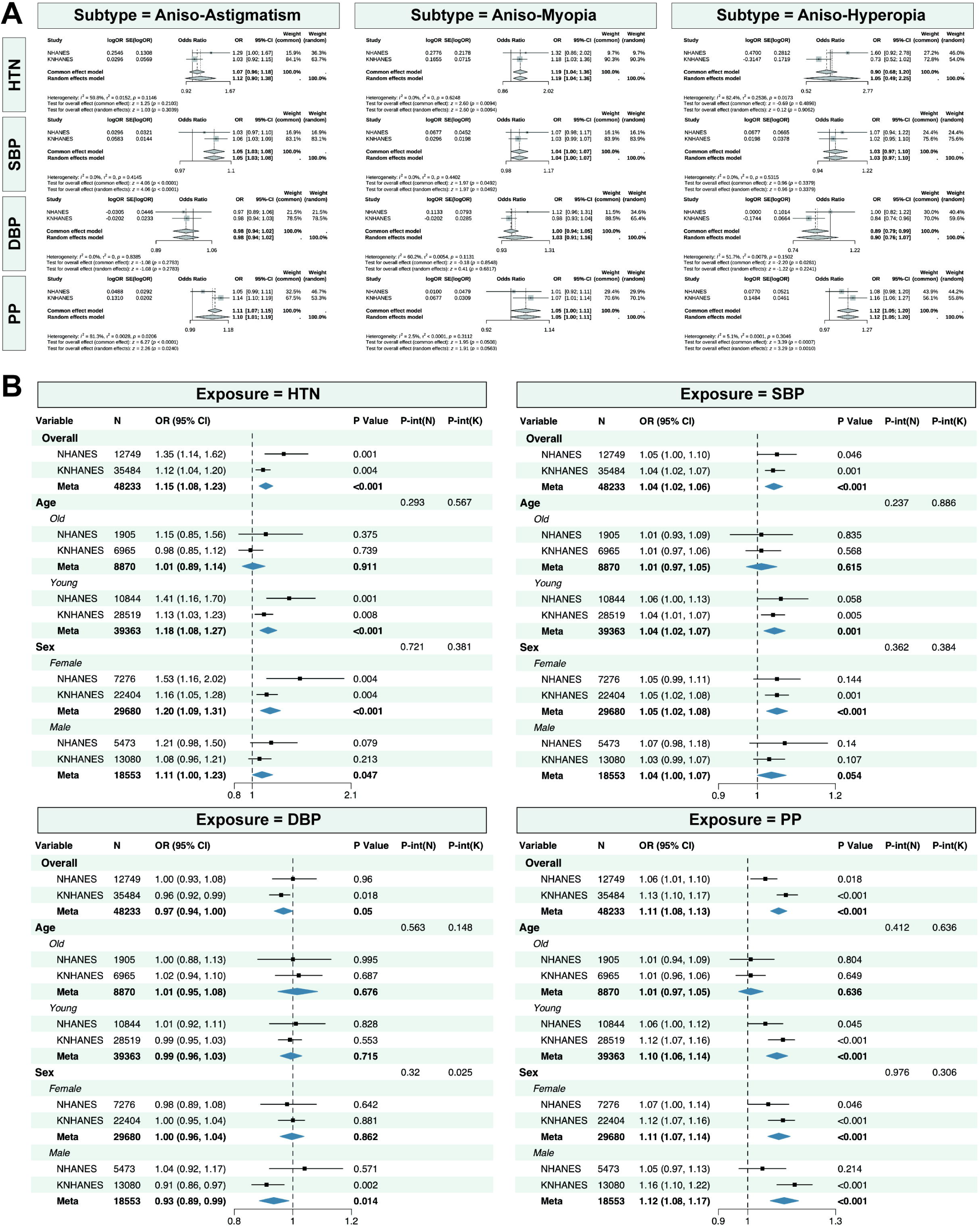
Subgroup analysis and subtype analysis for the association between the risk of anisometropia and hypertension as well as the association between the risk of hypertension and the subtype of anisometropia. (A) Subtype analysis. (B) Subgroup analysis.

As demographic variables might confound the association between the risk of anisometropia and hypertension, subgroup analyses were performed stratified by age and sex (Figure 3B). A significant association between hypertension, BPCs, and the incidence of anisometropia was observed in most subgroups. However, no substantial association was detected between hypertension and anisometropia in older individuals, between SBP and anisometropia in older males, DBP and anisometropia in younger females, or PP and anisometropia in older individuals.

These findings suggest that hypertension and its components may differentially contribute to the development of anisometropia across subtypes.

### 3.6. Mediation Analysis

Ocular structure is related to blood pressure and the development of refractive error^31,32^. To investigate the structural pathways linking blood pressure to anisometropia, we hypothesized that blood pressure may influence anisometropia via two patterns of interocular asymmetry. Further details can be found in the Methods section. Therefore, we conducted a mediation analysis on the OCT subcohort (N: 4,913–6,839).

Mediation analyses demonstrated that interocular CMT asymmetry mediated 4.11% and 4.93% of the associations of SBP and PP with anisomyopia, respectively. High DBP was significantly associated with a greater difference in interocular AL (path a, P = 0.049), which in turn was positively associated with anisohyperopia (path b, P < 0.001), yielding a significant indirect effect (P for ACME = 0.035), whose direction of the indirect effect was opposite to those of the overall association observed in the subtype association analyses (Figure 4A).

**Figure 4:**
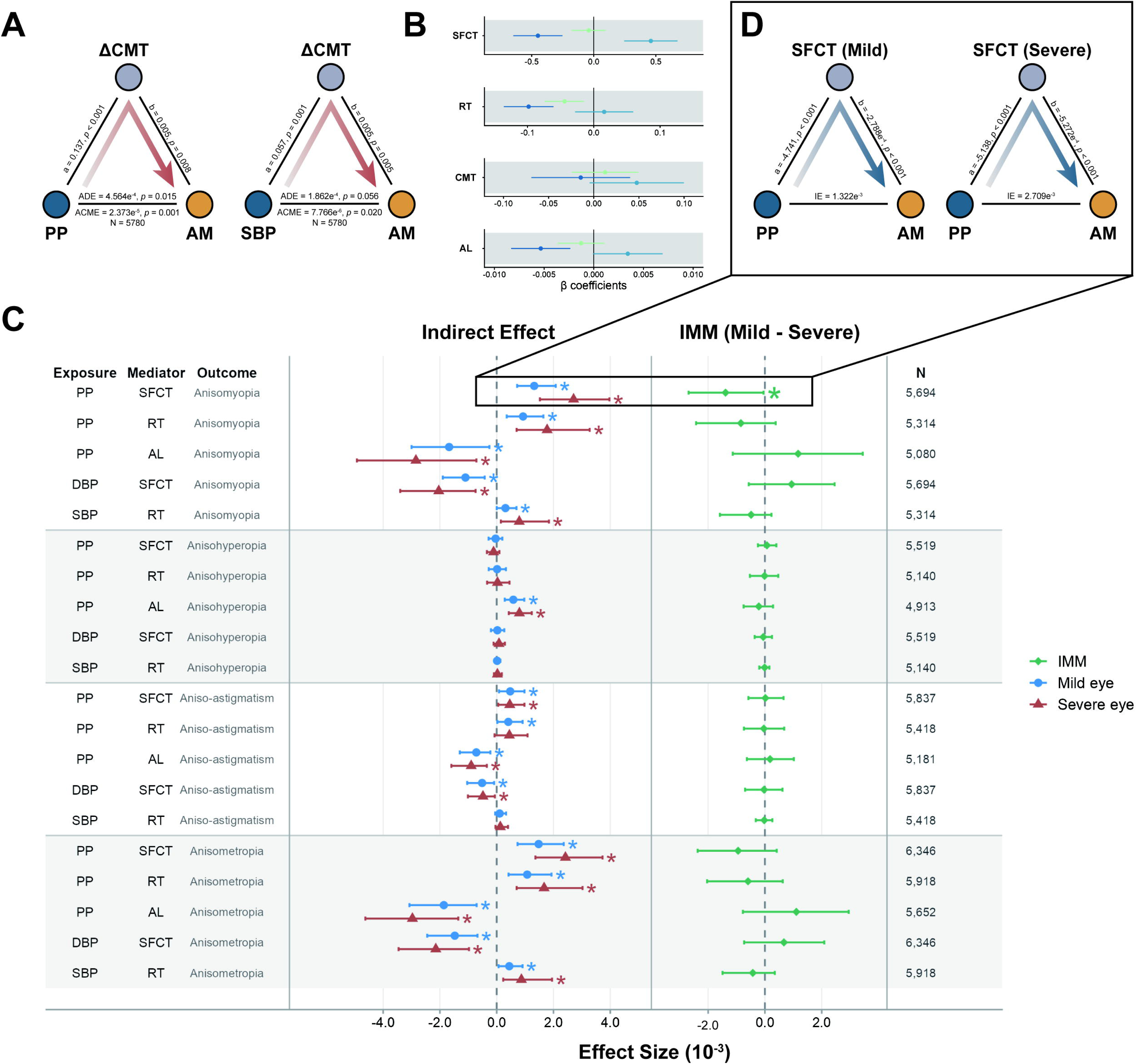
Mediation and moderated mediation analyses of ocular structural asymmetry in the associations between BPCs and anisometropia subtypes. (A) The path diagrams illustrate the mediating effects of interocular central macular thickness asymmetry (ΔCMT) in the associations of PP and SBP with anisomyopia. (B) Estimated coefficients from GEE models describing the associations between BPCs and OCT-derived ocular structural parameters. (C) The Forest plot summarizes the moderated mediation effects of OCT parameters in the severe and mild eyes on the associations between BPCs and anisometropia subtypes, estimated using the manymome framework. (D) The path diagrams show the significant moderated mediation models identified in panel C. Abbreviations: AL, axial length; SFCT, subfoveal choroidal thickness; RT, retinal thickness; CMT, central macular thickness. IMM, Index of Moderated Mediation.

Moreover, GEEs were used to quantify the effects of BPC on OCT parameters (Figure 4B). In multivariate GEE models, higher DBP and lower PP were independently associated with greater SFCT and longer AL (DBP-SFCT: coefficient, 0.459 μm/mmHg; 95% CI: 0.244–0.674□ μm/mmHg; P < 0.001; N = 13,126. PP-SFCT: coefficient, −0.450 μm/mmHg; 95% CI: −0.648 – −0.252□ μm/mmHg; P < 0.001; N = 13,126. PP-AL: coefficient, −0.005 mm/mmHg; 95% CI: −0.008– −0.002□mm/mmHg; P < 0.001; N = 11,706). Additionally, both higher SBP and higher PP were independently associated with thinner RT (SBP-RT: coefficient, −0.044 μm/mmHg; 95% CI: −0.074 – −0.014□ μm/mmHg; P = 0.004; N = 12,193. PP-RT: coefficient, −0.098 μm/mmHg; 95% CI: −0.136 – −0.061□ μm/mmHg; P < 0.001; N = 12,193).

As BPCs and ocular structures were found to be associated, moderated mediation analyses were performed to quantify the structural pathways linking blood pressure to anisometropia subtypes.

Figure 4C illustrates the conditional indirect effects of PP and other BPC on the risk of anisometropia, mediated by OCT parameters, stratified by eye severity. Across most exposure-mediator-outcome pathways, the Index of Moderated Mediation (IMM) was not statistically significant, suggesting that the indirect effects of BPC on anisometropia and its subtypes were comparable between the more-affected and less-affected eyes.

The anisomyopia subgroup showed a notable exception, where the indirect effect of PP on the risk of anisomyopia via SFCT was significantly stronger in the severe eye (IE_severe = 2.71e^-3^, 95% CI [1.52e^-3^, 3.97e^-3^]) than in the mild eye (IE_mild = 1.32e^-3^, 95% CI [7.21e^-4^, 2.08e^-3^]), with a significant IMM of −1.39e^-3^ (95% CI [-2.69e^-3^, −5.04e^-5^], P < 0.05) (Figure 4D). This finding suggests that higher PP differentially attenuates choroidal thickness in the more myopic eye, specifically among anisomyopic individuals, thereby contributing disproportionately to the development of interocular asymmetry.

### 3.7. Identification of hub genes that regulate hypertension and anisometropia

To identify hub genes linking hypertension and the progression of anisomyopia, the 467 time-interaction DEGs in anisomyopia underwent ClusterGVis analysis and were classified into six temporal clusters (Figure 5A). Cluster 5 (C5), which was most enriched for hypertension-related genes, decreased across the 24-h, 48-h, and 72-h time points, with pathway enrichment in Rho protein signal transduction, visual perception, and regulation of blood coagulation, suggesting progressive dysregulation of vascular and neurosensory processes during the development of anisomyopia.

**Figure 5:**
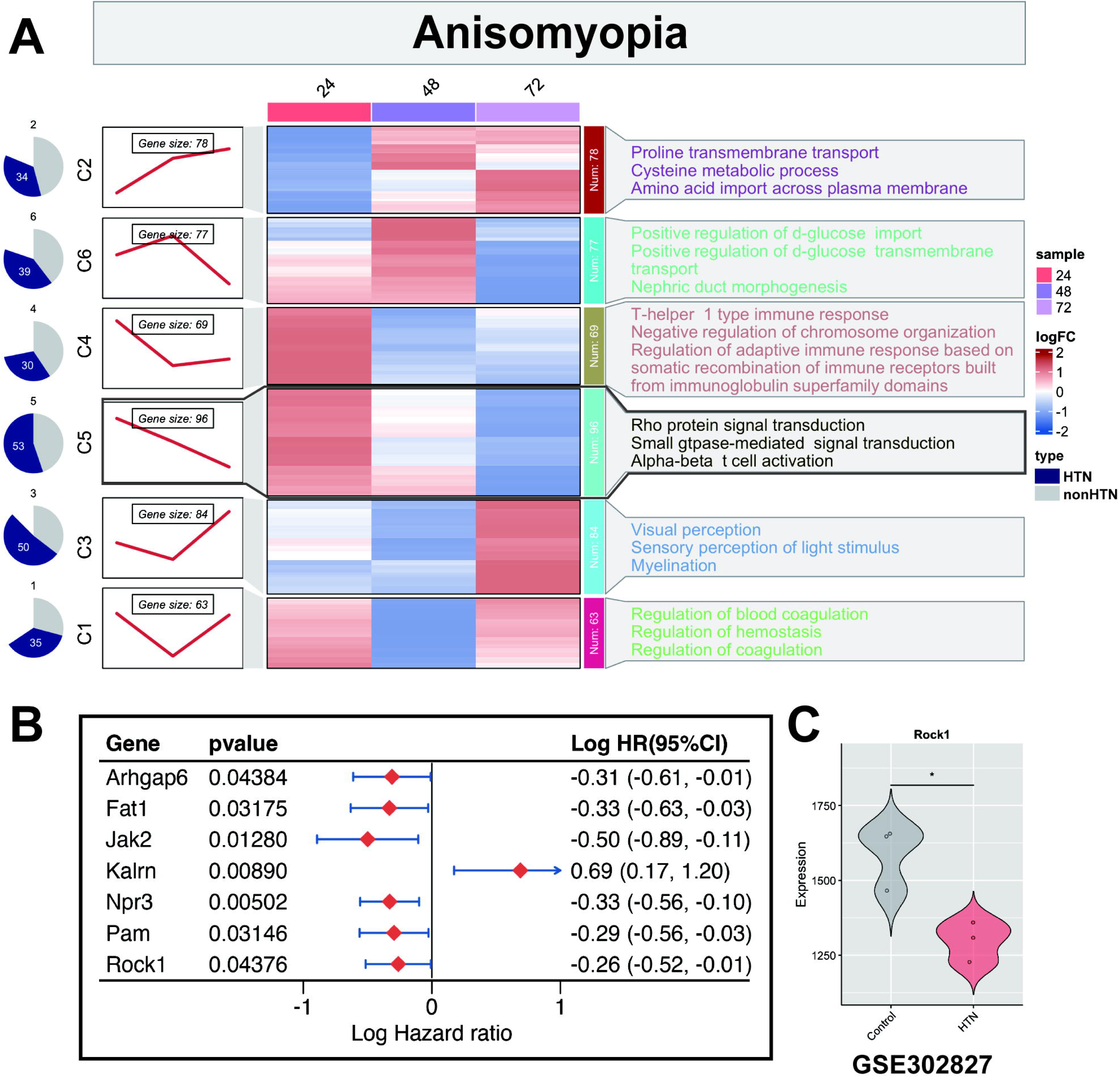
Identification of hub genes regulating the progression of anisomyopia under hypertensive conditions. (A) Temporal expression clusters of time-interaction DEGs in anisomyopia across 24, 48, and 72 h, with pathway enrichment annotations. (B) Forest plot of Cox regression results identifying prognostic markers in anisomyopia. (C) The violin plot shows the expression of Rock1 in SHR versus WKY control models (GSE302827); *P < 0.05.

Cox regression analysis was performed on GSE78042 and identified seven prognostic markers in anisomyopia (Figure 5B), among which Kalrn was a risk factor (HR > 1), while Arhgap6, Fat1, Jak2, Npr3, Pam, and Rock1 were protective factors (HR < 1).

To evaluate the expression of these prognostic markers under hypertensive conditions, gene expression levels were compared between SHR and WKY control models using GSE302827. Rock1, a key protective factor in the progression of anisomyopia, was significantly downregulated in the hypertensive group (P < 0.05; Figure 5C), suggesting that hypertension may impair this protective effect, thereby facilitating the development of anisomyopia.

## 4. Discussion

In this large-scale, multi-national population-based study, a consistent positive association was found between hypertension and anisometropia across two ethnically distinct cohorts, with the association partially mediated by interocular asymmetry in OCT structural parameters, including CMT and SFCT, between BPC and anisomyopia. Additionally, subtype analyses revealed that different BPCs differentially contribute to anisomyopia, aniso-astigmatism, and anisohyperopia. This is the first study to integrate mediation analysis with time-course transcriptomic profiling to elucidate the mechanism underlying hypertension-related anisometropia. The study identified ROCK1 as a candidate molecular target linking hypertension to the progression of anisomyopia.

Hypertension is correlated with a spectrum of ocular complications, including hypertensive retinopathy, choroidopathy, and optic neuropathy^33–34^. These conditions share a common pathophysiological basis that sustained increase in blood pressure induces progressive arteriolar remodeling and an increase in vascular stiffness, ultimately disrupting ocular perfusion and the blood-retina barrier^31^. SBP is influenced by large artery stiffness, wave reflections, and peripheral vascular resistance^35–37^, reflecting the hemodynamic load on the arterial system. PP, defined as the difference between SBP and DBP, serves as a surrogate marker of arterial stiffness^36–38^. In this study, SBP and PP, but not DBP, were positively associated with the risk of anisometropia, while the subgroup analyses suggested that the association between hypertension and anisometropia was significant in adults who were below 60 years old, as well as in females. This pattern of differential associations of BPCs with risk is consistent with the concept of wide PP hypertension^39^, a hemodynamic phenotype characterized by substantially higher systolic pressure and PP arising from large artery stiffening after midlife. Unlike mean arterial pressure-driven hypertension, wide PP hypertension generates pulsatile stress that may be transmitted to the microvasculature of the eye, potentially mediating differential structural remodeling between the two eyes. These findings suggest that the pulsatile stress rather than the steady-state hemodynamic burden may be the dominant driver of interocular asymmetry in hypertensive patients.

Ocular hemodynamics is closely associated with the development of refractive error^40–42^, and refractive development is associated with ocular microcirculation^43^. Insufficient choroidal blood flow perfusion can induce myopia in animal models^44^, while interocular differences in choroidal blood supply occur in myopic anisometropia^11^, and asymmetric optic nerve head blood flow occurs in hyperopic anisometropia^46^. These findings suggest that interocular hemodynamic asymmetry may be a structural substrate for the development of anisometropia, making anisometropia a hemodynamically-mediated form of hypertensive ocular disease, rather than purely an optical or developmental phenomenon. This study provided the first large-scale epidemiological evidence supporting this hypothesis. For anisometropia, hypertension was associated with a 19% increase in risk, and additional significant associations were observed for SBP and PP. For aniso-astigmatism, SBP was associated with a 5% increase in risk per 10 mmHg increase, and additional significant associations were observed for PP. For anisomyopia, each 10 mmHg increase in SBP was associated with a 4% increase in risk, whereas each 10 mmHg increase in PP was associated with a 5% increase in risk, although this association did not reach statistical significance (P = 0.051).. For anisohyperopia, each 10 mmHg increase in PP was associated with a 12% increase in risk, whereas each 10 mmHg increase in DBP was associated with a 12% decrease in risk.

Axial length, SFCT, RT, and CMT are key OCT-derived structural parameters reflecting the pathological changes in ocular structures, which are associated with the development of refractive error^47–52^. Sustained hypertension induces structural changes in the retinal microvasculature, including microvascular damage and narrowing of arterioles^53,54^, which serves as the pathological bridge between high blood pressure and downstream ocular structural remodeling. Hypertension is associated with changes in ocular structure^55,56^. However, the differential contributions of BPCs to these structural changes remain unclear. This study demonstrated that higher DBP and lower PP were independently associated with greater SFCT and longer AL, while higher SBP and higher PP were independently associated with thinner RT. This pattern agrees with the hypothesis that pulsatile hemodynamic stress, captured by SBP and PP, preferentially affects the retina through ischemic thinning, whereas steady-state perfusion pressure, captured by DBP, may sustain choroidal filling and axial elongation through distinct vasoactive mechanisms. These BPC-specific structural associations served as the framework for our subsequent subtype-stratified mediation analyses.

Different types of refractive error are supported by distinct structural mechanisms^32^, for which we examined how different ocular structures mediate the relationship between BPCs and anisometropia subtypes. As anisometropia is the phenotype of interocular divergence, we hypothesized that two parameters in the ocular structure within an individual may have different patterns mediating the relationship across different subtypes of anisometropia. As the structural profiles were subtype-specific, we hypothesized that BPCs may influence anisometropia through two distinct patterns of OCT-mediated asymmetry.

The first pattern involves BPC-driven absolute interocular differences in OCT parameters directly mediating anisometropia. Among the candidate BPC-□ OCT pathways derived from significant BPC–anisometropia associations, asymmetry in interocular CMT mediated the associations of SBP and PP with anisomyopia, accounting for 4.11% and 4.93% of the associations, respectively. However, interocular AL difference mediated the associations of DBP with anisohyperopia (P for ACME = 0.035), whose direction of indirect effect was opposite to those of the overall association observed in the subtype association analyses. This selectivity reflects two complementary properties of the central macular region. First, the fovea is metabolically the most demanding of any retinal region, making CMT highly sensitive to interocular differences in hemodynamic supply. Even subtle asymmetries in macular perfusion between the two eyes may manifest as detectable differences in CMT before other structural parameters diverge. Second, even small changes in retinal and choroidal thickness have disproportionately large downstream consequences for scleral remodeling and axial elongation, as the choroid serves as a conduit for retina-to-sclera signaling molecules and a regulator of scleral oxygenation^57^. A modest difference in interocular CMT, amplified through this retina–choroid–sclera cascade, may be able to generate measurable anisomyopia. The anatomical basis for choroidal laterality may further augment this asymmetry through two mechanisms: vascular anatomical asymmetry, whereby the shorter pathway of the right carotid system may enhance choroidal perfusion and ocular dominance effects in the right eye, as approximately 67% of individuals exhibit right-eye dominance, with the dominant eye modulating local choroidal thickness through metabolic demand^58^.

The second pattern involves BPC-driven interocular differences in OCT parameters directly mediating anisometropia, whereby interocular asymmetry occurs due to differential structural susceptibility rather than absolute differences in OCT parameters between the two eyes. By conducting moderated mediation analyses, we found that the IMM was not statistically significant across most exposure-mediator-outcome pathways, which indicated that the indirect effects of BPC on anisometropia and its subtypes were generally comparable between the severe and mild eyes. The anisomyopia subgroup was an exception, where the indirect effect of PP on anisomyopia through SFCT was significantly stronger in the severe eye than in the mild eye. This finding suggests that high PP may preferentially influence choroidal structural alterations in the more-affected eye, thereby enhancing interocular asymmetry and promoting the development of anisomyopia. The stronger mediation effect may reflect the greater cumulative hemodynamic burden borne by the severe eye. As these eyes have undergone extensive structural remodeling, they may exhibit greater microvascular vulnerability and lower autoregulatory capacity in response to a sustained increase in blood pressure, making them more susceptible to PP-related choroidal changes and their downstream effects on ocular growth^58^.

At the molecular level, this study identifies ROCK1 as a key link between hypertension and anisomyopia. ROCK1 is a downstream effector of RhoA signaling that encodes a serine/threonine protein kinase that is activated by RhoA binding^59,60^. RhoA/ROCK signaling is a strong mediator of hypertension pathophysiology and contributes to vascular smooth muscle contraction, endothelial dysfunction, and vascular remodeling^61^. In contrast to the commonly reported activation of ROCK signaling in hypertensive vascular tissues, the analysis in this study identified a reduction in the expression of ROCK1 in the aorta tissues of spontaneously hypertensive rats after six weeks, probably reflecting a maladaptive compensatory response to sustained hypertensive stress. Consistent with this, the expression of ROCK1 was progressively reduced throughout the course of development of anisomyopia, suggesting impaired ROCK1-mediated vascular and cytoskeletal regulation as a contributor to asymmetric ocular growth. As Rho/ROCK signaling plays a role in cytoskeletal organization and ocular biomechanics^62,63^, its chronic downregulation may impair remodeling of the extracellular matrix and microvascular perfusion in the posterior segment, contributing to the asymmetric changes in choroidal and CMT observed between severe and mild anisomyopic eyes. A decrease in ROCK1-mediated actomyosin contractility may compromise the integrity of choroidal stroma and vascular autoregulation, leading to differential choroidal thinning or thickening across eyes. Simultaneously, disrupted cytoskeletal homeostasis in retinal cells under chronic hypertensive stress may render the retina vulnerable to thickness asymmetry, particularly in the context of impaired local perfusion driven by the suppression of ROCK1. In severe anisomyopic eyes, the expression of ROCK1 was transiently elevated at the early induction stage before decreasing in later phases. This biphasic pattern may be explained by caspase-3-mediated cleavage of ROCK1 during the execution phase of apoptosis, which is significantly higher in myopic retinal tissues^64^. Caspase-3 cleaves the auto-inhibitory C-terminal domain of ROCK1, resulting in its constitutive activation^65^. This transient, apoptosis-driven increase in the activity of ROCK1 in the early stage of severe myopia may represent a distinct regulatory mechanism superimposed on the broader hypertension-associated suppression of the expression of ROCK1. Inhibition of RhoA/ROCK signaling can suppress choroidal neovascularization and attenuate vascular remodeling^66,67^. In the context of chronic downregulation of ROCK1 observed in our model, reduced ROCK-mediated vascular tone may modulate choroidal perfusion and tissue compliance, thereby contributing to the choroidal thickening and alterations in the extracellular matrix observed in the posterior segment. These findings suggest that dysregulation of the RhoA/ROCK1 axis, driven by hypertensive stress and intensified by apoptotic activation in severe myopic eyes, may serve as a convergent molecular pathway underlying the development of hypertension-associated anisomyopia.

A methodological strength of this study is the application of the DML framework, which overcomes the limitations of conventional regression approaches when dealing with high-dimensional and non-linear confounders. The consistency of ATE estimates across multiple machine learning algorithms and both cohorts further supports the robustness of the findings of this study.

This study, however, had several limitations. First, the cross-sectional design precludes causal inference; the temporal relationship between the onset of hypertension and the development of anisometropia could not be established. Second, OCT parameter measurements were available only in the KNHANES cohort, which restricted the generalizability of the mediation findings. Third, residual confounding from unmeasured variables, such as genetic factors, could not be excluded. Fourth, the transcriptomic analyses were based on animal models, and the translational relevance to human anisometropia requires validation.

## Data Availability

All data produced in the present study are available upon reasonable request to the authors

## 5. Acknowledgements

We sincerely thank the staff and participants of the NHANES and KNHANES studies for their valuable contributions. This work was supported by the Research Project on Chronic Disease Management in 2025 (Grant No. GWJJMB202510025142).

