## Supplementary figures and images for "Hypertension-Associated Anisometropia: Interocular Choroidal and Macular Asymmetry as Potential Mediators"

### Supplemental FigureS 1

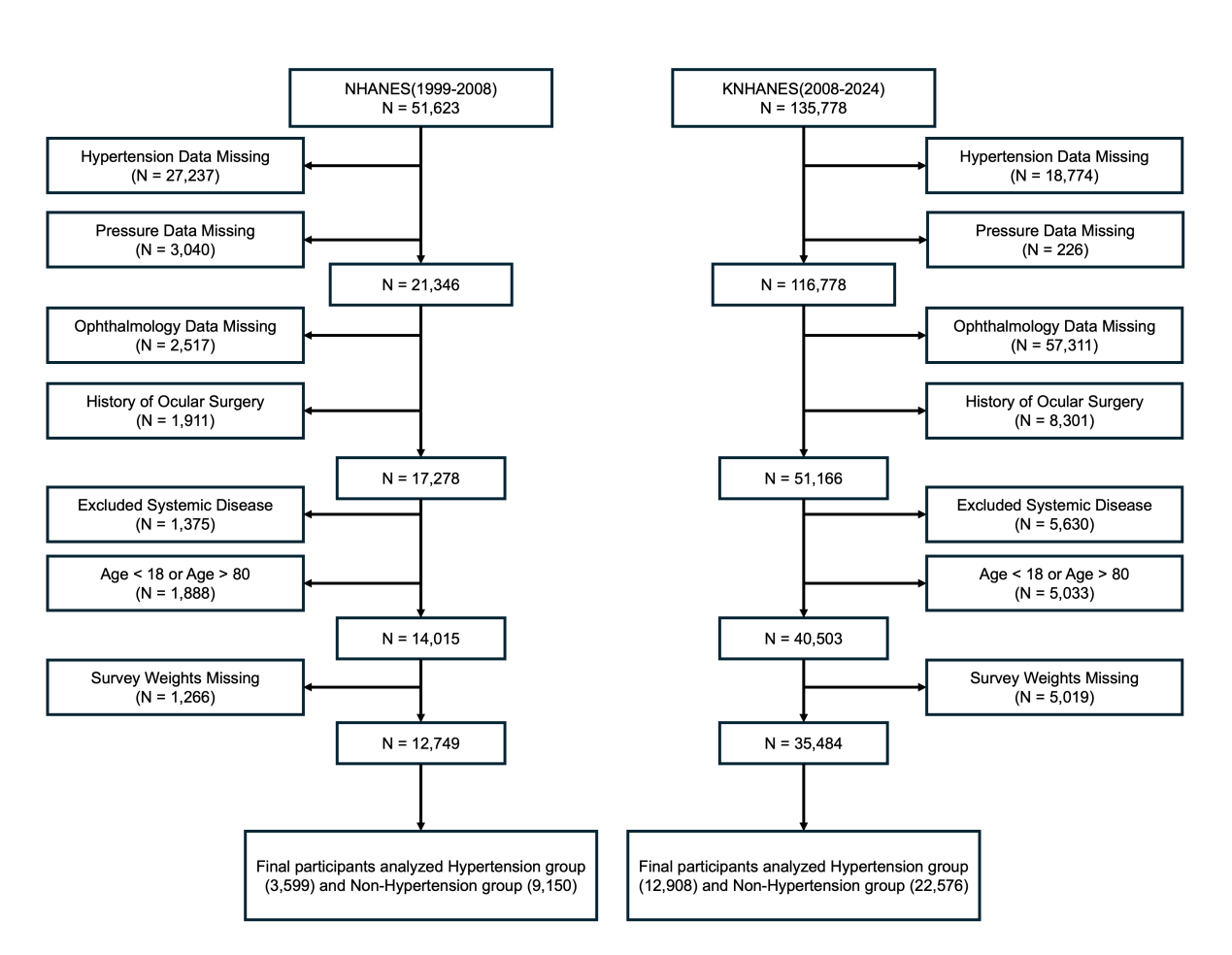

FigureS 1: Flowchart of the study participants.
